# Evidence for a shared genetic contribution to loneliness and Borderline Personality Disorder

**DOI:** 10.1101/2023.03.16.23286984

**Authors:** Anna Schulze, Fabian Streit, Lea Zillich, Swapnil Awasthi, Alisha S M Hall, Martin Jungkunz, Nikolaus Kleindienst, Josef Frank, Cornelia E Schwarze, Norbert Dahmen, Björn H Schott, Markus Nöthen, Arian Mobascher, Dan Rujescu, Klaus Lieb, Stefan Roepke, Sabine C Herpertz, Christian Schmahl, Martin Bohus, Stephan Ripke, Marcella Rietschel, Stefanie Lis, Stephanie Witt

## Abstract

Loneliness, influenced by genetic and environmental factors such as childhood maltreatment, is one aspect of interpersonal dysfunction in Borderline Personality Disorder (BPD). Numerous studies link loneliness and BPD and twin studies indicate a genetic contribution to this association. The aim of our study was to investigate whether genetic predisposition for loneliness and BPD risk overlap and whether genetic risk for loneliness contributes to higher loneliness reported by BPD patients, using genome-wide genotype data. We assessed the genetic correlation of genome-wide association studies (GWAS) of loneliness and BPD using linkage disequilibrium score regression and tested whether a polygenic score for loneliness (loneliness-PGS) was associated with case-control status in two independent genotyped samples of BPD patients and healthy controls (HC; Witt2017-sample: 998 BPD, 1545 HC; KFO-sample: 187 BPD, 261 HC). In the KFO-sample, we examined associations of loneliness-PGS with reported loneliness, and whether the loneliness-PGS influenced the association between childhood maltreatment and loneliness. We found a genetic correlation between the GWAS of loneliness and BPD in the Witt2017-sample (rg = .23, *p* = .015), a positive association of loneliness–PGS with BPD case-control status (Witt2017-sample: NkR² = 2.3%, *p* = 2.7*10^-12^; KFO-sample: NkR² = 6.6%, *p* = 4.4*10^-6^), and a positive association between loneliness-PGS and loneliness across patient and control groups in the KFO-sample (*β* = .185, *p* = .002). The loneliness-PGS did not moderate the association between childhood maltreatment and loneliness in BPD. Our study is the first to use genome-wide genotype data to show that the genetic factors underlying variation in loneliness in the general population and the risk for BPD overlap. The loneliness-PGS was associated with reported loneliness. Further research is needed to investigate which genetic mechanisms and pathways are involved in this association and whether a genetic predisposition for loneliness contributes to BPD risk.

## 1 Introduction

A pervasive feeling of loneliness is one aspect of interpersonal dysfunction in Borderline Personality Disorder (BPD) [1]. It is defined as a negative affective state resulting from the discrepancy between desired and experienced social connectedness [2]. Objective social isolation may contribute to feelings of loneliness, but it is neither necessary nor sufficient to fully explain them: for example, people who are embedded in a large social network may feel lonely, whereas people with a small number of social contacts may not [3]. A short-lasting acute experience of loneliness is assumed to have a beneficial evolutionary function, promoting behaviours to reconnect to the social environment [4]. In contrast, long-lasting feelings of loneliness have been linked to an increased risk to health and a detrimental effect on the course of both somatic and mental disorders [5–7]. It has been discussed for a long time that loneliness is partly attributable to environmental factors like childhood maltreatment as well as to genetic factors, which is supported by the results of family and twin studies [8–10]. In the recent years, genome-wide association studies (GWAS) have identified genetic variation associated with loneliness [11] and, to a lesser degree, with BPD [12]. Despite findings from twin studies supporting a genetic correlation between BPD and loneliness, it has not yet been investigated whether the genetic variants associated with loneliness are more common in individuals with BPD. Therefore, the current study uses genome-wide genetic data to assess the genetic and phenotypic overlap between loneliness and BPD and explore its association with childhood maltreatment.

While loneliness is a transdiagnostic feature of psychopathology, it plays a central role in interpersonal dysfunction in BPD: individuals with BPD often report a lack of sense of belonging and the fear of being abandoned or socially excluded [13]. BPD is a personality disorder with a prevalence of 0.92–1.90 % in western countries [14], associated with a high economic burden to the health care system and economy [e.g. 15] and partly attributable to genetic factors [12]. Adler and Buie observed intensely painful aloneness as a core experiential state in their characterological work with BPD patients already in 1979 [16], Gunderson emphasized that the fear of aloneness discriminates BPD from other personality disorders [1] and contributes to their high sensitivity towards social rejection [17, 18]. Additionally, the interpersonal style of BPD patients, characterized as intense and unstable, maintains the pattern of recurrent interpersonal problems [17]. Several studies have shown increased levels of loneliness in BPD, which have been linked to smaller social networks [19, 20]. Furthermore, loneliness in BPD is linked to impairments of social-cognitive processing such as the experienced confidence in one’s own social-emotional judgments [21] and the strength of basic affiliative behaviors, such as behavioral mimicry [22]. BPD patients describe the feeling of loneliness as a persisting state arising as early as in childhood [23], suggesting that an increased propensity towards loneliness might be apparent at an early age already. Disorder-specific therapeutic interventions are successful in improving acute symptoms such as impulsivity or non-suicidal self-harming behaviours, but are less effective in reducing the feeling of loneliness with consequences for persistence of impairments in the patients’ social functioning level [24, 25]. Therefore, a deeper understanding of the determinants of loneliness in BPD is of particular interest.

Genetic studies have contributed substantially to our understanding of inter-individual differences in mental health [26]. Twin and family studies aim to estimate the influence of genetic and environmental influences on the variation of traits or disorder risk using the information on genetic relatedness and shared family environment [27]. As a complementary approach, genome-wide association studies (GWAS) aim to identify single nucleotide polymorphisms (SNPs), i.e. common changes of single base pairs in the DNA, associated with a specific phenotype. For psychiatric symptoms and disorders, the so called SNP-based heritability, i.e. the variance explained by the common variants assessed in a GWAS, usually accounts for around one third of the heritabilities estimated in twin studies [26]. Besides insights into specific genes and pathways involved in disease etiology, GWAS also allow the estimation to what degree the association signal, and thereby the underlying genetic factors, are shared between disorders and traits. For example, genetic correlations are point estimates of the genetic similarity, and can be estimated using summary statistics of independent GWAS with linkage disequilibrium (LD)-score regression [28]. Another approach is the calculation of polygenic scores (PGS) based on the identified associations in GWAS (discovery samples) in independent target samples of healthy or affected individuals, representing the individual’s propensity towards a disease or trait [29]. PGS of psychiatric phenotypes still only explain a limited amount of variance and are therefore not applicable in clinical practice. However, they have the advantage that they can be also computed in smaller samples, and have proven to be a useful tool in research to investigate, for example, the association of the genetic predisposition to a trait with related phenotypes.

Family and twin studies demonstrate that genetic factors contribute to BPD as well as to loneliness. The heritability of BPD is estimated to be around 46–69% [30, 31], while genetic factors explain approximately 38–48% of the variance in loneliness in adults [8–10]. Analyses of shared genetic and environmental factors for borderline personality features and loneliness revealed a high genetic correlation of *r* = .64, but also a unique environmental correlation of *r* = .40 in a twin study [32]. Findings of another twin study indicated that loneliness might mainly be a consequence of the genetic determinants of BPD traits [33].

A GWAS assessing borderline personality features as a dimensional trait found a SNP-heritability of 23% [34]. Moreover, the polygenic score (PGS) for borderline personality features based on this GWAS was found to have a positive association with neuroticism [35], a personality trait associated with loneliness [36]. So far, one case-control GWAS, i.e. comparing BPD patients diagnosed using established diagnostic systems to controls, has been performed which did not identify associated single variants but indicated significant gene-based associations in the genes coding for dihydropyrimidine dehydrogenase (*DPYD*) and plakophilin-4 (*PKP4*), which have been previously linked to other mental disorders, e.g. schizophrenia [12]. BPD was found to have positive genetic correlations with major depression, bipolar disorder, and schizophrenia [12] as well as with the personality traits neuroticism and, to a lesser degree, openness to experience [37]. At the same time, a recent GWAS in the UK Biobank identified 15 genome-wide significant loci associated with loneliness [11], measured by three variables assessing the feeling of loneliness, the frequency of interacting with others and the possibility to confide in others. In a phenome-wide association study the PGS for loneliness was associated with personality traits, especially neuroticism, and a wide range of somatic but also psychiatric disorders such as mood disorders and depression [38]. However, this approach has not been used yet to study the association of loneliness with BPD.

Childhood maltreatment has been identified a major environmental risk factor for BPD with individuals with a diagnosis of BPD being around thirteen times more likely to report childhood maltreatment than non-clinical controls [39, 40]. Some studies suggest an interaction between adverse life events and the genetic risk for mental disorders [e.g. 41, 42]. For example, the genetic correlation of major depression disorder with waist circumference was significantly greater in individuals reporting exposure to trauma compared to those not reporting trauma exposure [41]. Moreover, genetic vulnerability and stressful life events have not only shown additive but also an interactive effect on depressive symptoms with especially increased depression scores in subjects with both, stressfull life events and high polygenic risk [42]. Since childhood maltreatment is also related to a higher risk for perceived social isolation in adulthood [43–45], childhood maltreatment might be crucial in gene-environment interactions associated with loneliness in BPD.

The aim of the present study was to investigate the genetic and phenotypic overlap between loneliness and BPD and explore its association with childhood maltreatment. For this, we analyzed data from two independent genotyped BPD samples. The first, previously published, sample (Witt2017-sample) consisted of 998 BPD patients and 1545 HCs [12]. The second sample (KFO-sample) was an independent clinical sample of 187 well-characterized patients with BPD and 261 HCs who provided data on loneliness and childhood maltreatment and were part of the sample recruited by the clinical research unit KFO 256 [46]. Due to the genetic correlation of loneliness with borderline personality features observed in a twin study [32] and known association of loneliness-PGS with psychiatric disorders in a phenomewide association study [38], we wanted to test this association in our samples in a first step. We expected 1) a positive genetic correlation between loneliness and BPD, 2) higher loneliness-PGS in BPD cases compared to controls, and 3) a positive association of the loneliness-PGS and an individual’s loneliness. Furthermore, we analyzed whether our findings can be explained by the genetic disposition to neuroticism, a personality trait associated with loneliness and BPD in the past [36–38]. Finally, we explored whether 4) the severity of childhood maltreatment predicts loneliness stronger for BPD patients with a high genetic risk for loneliness in the KFO-sample.

## 2 Materials and Methods

### 2.1 Witt2017-Sample – characteristics and methods

#### 2.1.1 Sample Characteristics

The Witt2017-sample consisted of a BPD GWAS sample described in detail previously in Witt et al. [12]. Briefly, controls and subjects meeting DSM-IV criteria for BPD were recruited at three university hospitals in Germany. All subjects provided written informed consent, and the study was approved by the local ethics committees. After quality control (see below), the sample consisted of 998 cases (91.58% female, mean age 29.58, range: 18–65 years, *SD* = 8.64) and 1545 controls (56.18% female, mean age 44.19 years, range: 18–72 years, *SD* = 13.24).

#### 2.1.2 Genetic correlation analysis

To obtain a point estimate of the genetic correlation of loneliness with BPD, we used LD-score regression [28]. LD-score regression allows the calculation of genetic correlations of GWAS that have been carried out in independent samples. Calculations were carried out with a free intercept and the European ancestry samples from the 1000 Genomes data as LD structure reference panel [47]. Summary statistics from the GWAS of loneliness (N = 445 024) [11] and the GWAS of BPD (998 cases, 1545 controls) [12] were used as input. In order to capture associations with the feeling of loneliness, as assessed with the ULS-R in the current study, we used the GWAS based on the single item ‘Do you often feel lonely?’ instead of the 3 item measure reflecting more strongly social isolation, that is, the frequency of social interaction.

#### 2.1.3 Polygenic Scores

For the present analyses, PGS were calculated based on an updated quality control and imputation procedure, which has been described in detail in Streit et al. [37]. Subjects were genotyped using Illumina Infinium PsychArray-24 Bead Chips (Illumina, San Diego, CA, USA). Genetic markers and subjects were filtered after the following exclusion criteria: genotypic and individual missingness (> 2%), missingness differences between cases and controls (> 2%), deviation from autosomal heterozygosity (|Fhet| > 0.2) or deviation from Hardy-Weinberg equilibrium (controls: p < 1*10-6, cases: p < 1*10-10). Additionally, subjects were excluded when they showed sex mismatches, cryptical relatedness, or were genetic outliers.

Imputation was performed with the publicly available reference panel from the Haplotype Reference Consortium (EGAD00001002729), using EAGLE/MINIMAC3 [default settings, variable chunk size of 132 genomic chunks; 48, 49], and best-guess genotypes were used for PGS analyses.

For PGS calculation, variants in the Witt2017-sample were filtered for imputation quality with an INFO score of >=0.9, and minor allele frequency of >=5%. PGS were then calculated using PRSice 2.1.6 clumping SNPs based on the p-value in the discovery samples and LD-structure in the target sample with standard settings (distance = 250kb, p = 1, r² = 0.1) [50]. PGS were calculated for loneliness (loneliness-PGS) using summary statistics from Day et al. [11] based on the GWAS for loneliness assessed as a single item, and for neuroticism using summary statistics from Nagel et al. [51], excluding SNPs with an INFO score <0.9 in the discovery sample. PGS were calculated for 10 p-value thresholds (p-value threshold (PT): 5*10−8, 1*10−6, 1*10−4, 0.001, 0.01, 0.05, 0.1, 0.2, 0.5 and 1.0; see Supplementary Table S1 for number of included SNPs). There was no sample overlap of the discovery samples with the Witt2017-sample.

PRSice2 [50] was used to calculate logistic regression models with case-control status as the dependent variable, and loneliness-PGS as a predictor of interest and the first 5 ancestry principal components (PC1–PC5) as covariates. As effect size measure, the increase in Nagelkerke’s pseudo-R² (NkR²), was calculated, comparing the full model (including PGS and covariates as predictors) to the reduced model (including only covariates as predictors).

### 2.2 KFO-Sample – characteristics and methods

#### 2.2.1 Sample Characteristics

A total of 448 female adult individuals who passed genetic quality control were included in the present analysis, 187 of whom met DSM-IV criteria for BPD (age *M* = 29.35, *SD* = 7.69) and 261 were healthy controls (HC, age *M* = 27.56, *SD* = 6.94). Participants of the BPD group were slightly older (*t* = –2.58, *p* = .010, *d* = –.25). Data on childhood traumatization was available for subsample of 409 participants (169 BPD, 240 HC), and data on loneliness for 290 individuals (155 BPD, 135 HC). For 276 subjects (144 BPD, 132 HC), both were available. Recruitment was carried out by the central project of the KFO 256, which is a clinical research unit funded by the German Research Foundation (DFG) dedicated to investigating mechanisms of disturbed emotion processing in BPD [46]. The diagnosis of BPD according to DSM-IV was made by trained clinical psychologists using the International Personality Disorder Examination [52], a semistructured clinical interview assessing personality disorders for both the DSM-IV and the ICD-10 classification systems. All patients met at least five of the nine DSM-IV criteria for BPD.

General exclusion criteria were a lifetime history of psychotic or bipolar I disorder, current substance addiction, current pregnancy, history of organic brain disease, skull or brain damage, severe neurological illness or psychotropic medication at the time of the testing as well as a positive urine toxicology screen for illicit drugs. Additional exclusion criteria for the healthy controls were any lifetime or current psychiatric diagnoses.

The study was conducted in accordance with the Declaration of Helsinki and was approved by the Research Ethics Board of the University of Heidelberg. Subjects provided written informed consent prior to study participation.

#### 2.2.2 Measures

**Loneliness.** Loneliness, that is the subjective experience of social isolation, was assessed using the Revised University of California Los Angeles Loneliness Scale (ULS-R) [53, German version: 54]. The ULS-R consists of 20 items that are rated on a 5-point Likert scale (range: 1 ‘*not at all’* to 5 ‘*totally’*) combined in a sum score (range: 20–100) with higher scores indicating higher levels of loneliness. Internal consistency for the ULS-R was α = .972 (BPD: Cronbach’s α = .938; HCs: Cronbach’s α = .886).

**Childhood Matreatment.** Severity of childhood maltreatment was assessed using the short form of the Childhood Trauma Questionnaire (CTQ-SF) [55, German version: 56]. Subjects rated the frequency of maltreatment in childhood and adolescence in 25 items using a 5-point Likert scale (range: 1 ‘not at all’ to 5 ‘very often), with sum-scores ranging from 25 to 125. Internal consistency for the CTQ-SF was α = .954 (BPD: Cronbach’s α = .930; HCs: Cronbach’s α = .873).

#### 2.2.3 Genotyping, quality control and imputation

DNA was extracted from peripheral blood samples using automated DNA extraction with the chemagic Magnetic Separation Module I (Chemagen Biopolymer-Technologie, Baesweiler, Germany). All samples were genotyped using Illumina InfiniumGlobal Screening Arrays (Illumina, San Diego, CA, USA).

Genetic quality control and imputation for the KFO-sample was carried out in the frame of a larger ongoing BPD GWAS study [57], and was done as described for the Witt2017-sample. For the present analyses, the subjects from the KFO-sample were extracted from the larger data set, and homogeneity of the dataset was ensured by excluding subjects >|4.5| SD on the first 20 PCs.

#### 2.2.4 Polygenic Scores

PGS for loneliness and neuroticism were calculated and tested for association with case-control status using PRSice2 as described for the Witt2017-sample. There was no sample overlap of the discovery samples with the KFO-sample. The number of included SNPs for each p-value threshold are shown in Supplementary Table S1.

#### 2.2.5 Statistical Analysis

The loneliness-PGS which showed the strongest association with case-control status in sample 2, PT = 0.1, was selected for further analyses in the sample. To analyze the relationship between self-reported loneliness and the loneliness-PGS, we applied multiple linear regression analysis with the loneliness-PGS as predictor and ULS-R score as dependent variable, controlling for the target cohort’s specific principal components (PC1–PC5). To examine whether the loneliness-PGS moderates the association of the severity of childhood maltreatment and the ULS-R score in the BPD group, a moderation analysis with the z-standardized predictors CTQ score, ULS-R score and their interaction term was performed. We used the PROCESS macro by Hayes AF [58], which uses ordinary least squares regression, yielding unstandardized coefficients for all effects. Bootstrapping with 5000 samples together with heteroscedasticity consistent standard errors (HC3) [59] were employed to compute the confidence intervals. In all analyses, the first five PCs were included as covariates to control for population stratification. In addition, the PGS for neuroticism (neuroticism-PGS) was included as a covariate in a second step in order to control for its often reported association to BPD and loneliness [60].

## 3 Results

### 3.1 Genetic correlation

In the LD-score regression analysis, the BPD GWAS [12] showed a positive genetic correlation with the loneliness GWAS ([11]; rg = .23 [95% CI: .046 – .42]; *p* = .015).

### 3.2 PGS association with case-control status

Loneliness-PGS showed a positive association with BPD case-control status: higher loneliness-PGS were observed in the BPD cases (see Figure 1, details see Supplementary Tables S2-S7). For the Witt2017-sample, the strongest association was observed for the PT = 0.5 (*NkR*² = 2.3%, *p* = 2.7*10^-12^), and the association was replicated in the KFO-sample (PT = 0.1, *NkR*² = 6.6%, *p* = 4.4*10^-6^). When adding the best fit neuroticism-PGS (both studies PT = 0.1) as a covariate, a reduced *NkR²* was observed, but the association remained significant (Witt2017-sample: PT = 0.5, *NkR*² = 0.6%, *p* = 0.00019; KFO-sample: PT = 0.1, *NkR*² = 2.7%, *p* = 0.0021).

**Fig. 1:**
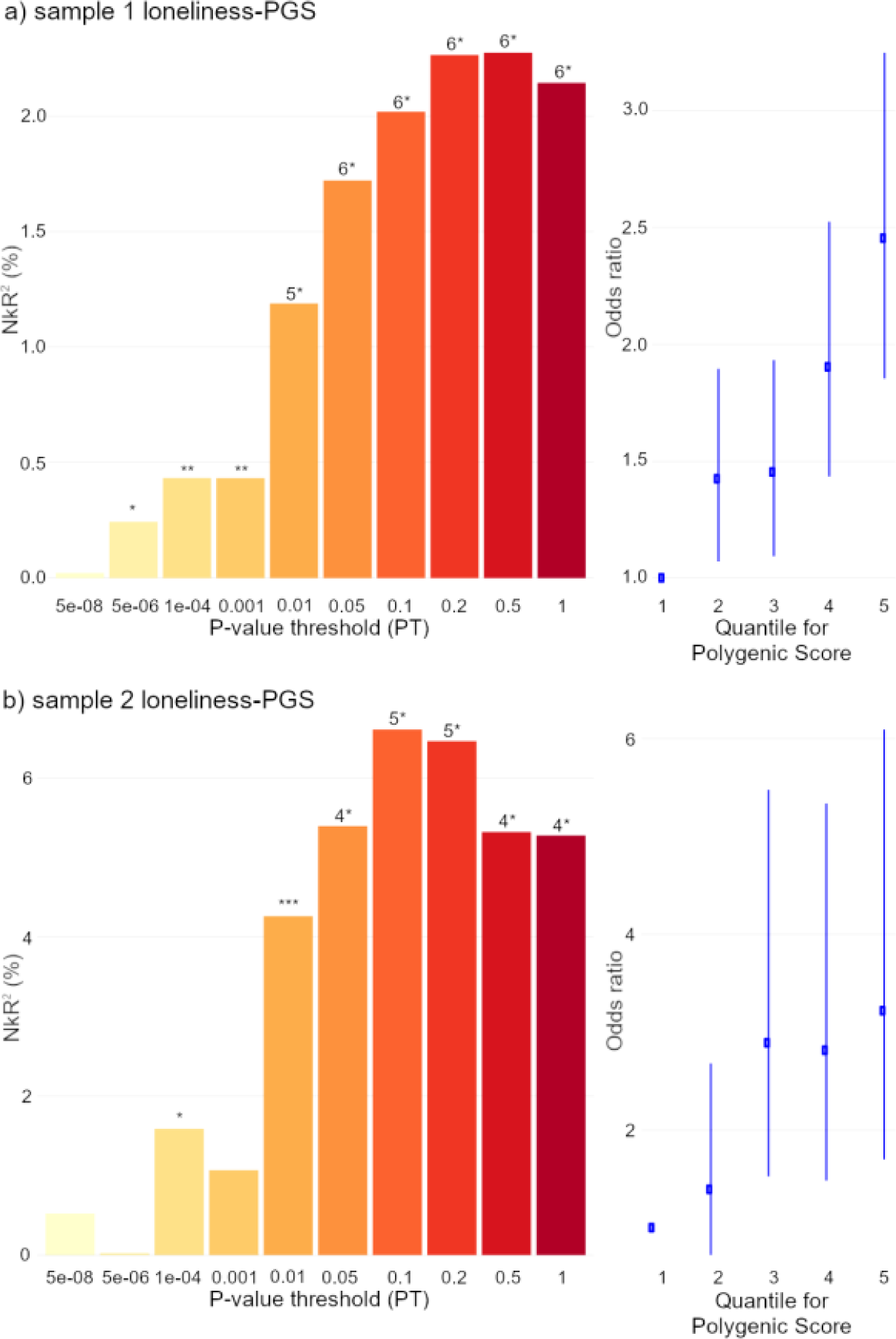
*Association of Loneliness Polygenic Scores (loneliness-PGS) with Borderline Personality Disorder case-control status.* Left panel: Nagelkerke’s R² describing explained variance in case-control status by PGS at ten P-value thresholds. Right panel: Odds ratio for case-control status depicted by loneliness-PGS quintile, with the first quintile as reference, depicted for the most strongly associated PT. Loneliness-PGS was based on Day et a. (2017). The number of SNPs included in the PGS are shown in Table 1. * *p* < 0.05; ** *p* < 0.005; *** *p* < 0.001; 4* *p* < 1×10−4; 5* *p* < 1×10−5; 6* *p* < 1×10−8

### 3.3 Prediction of loneliness by loneliness-PGS

BPD patients reported a higher level of loneliness than HC (*t* = –22.186, *p* < .001, *d* = –2.493). Multiple linear regression analyses revealed that the loneliness-PGS and PCs predicted 5.8% of the variance of the ULS-R score in the KFO-sample combined for patient and control groups (*F*(6, 283) = 2.92, *p* = .009, adjusted *R^2^* = .038), with the loneliness-PGS as the only significant predictor (*β* = .185, *p* = .002, Figure 2). This finding remained significant when we additionally controlled for the neuroticism-PGS (*F*(7, 282) = 3.46, *p* = .001, adjusted *R^2^* = .056, *β* = .126, *p* = .046). Separate analyses for the BPD and HC group revealed no significant relationship within the subgroups (BPD: *F*(6, 148) = 1.24, *p* = .287; HC: *F*(6, 128) = 1.05, *p* = .396, additionally controlled for neuroticism-PGS BPD: *F*(7, 147) = 1.06, *p* = .390; HC: *F*(7, 127) = 0.89, *p* = .513).

**Fig. 2.**
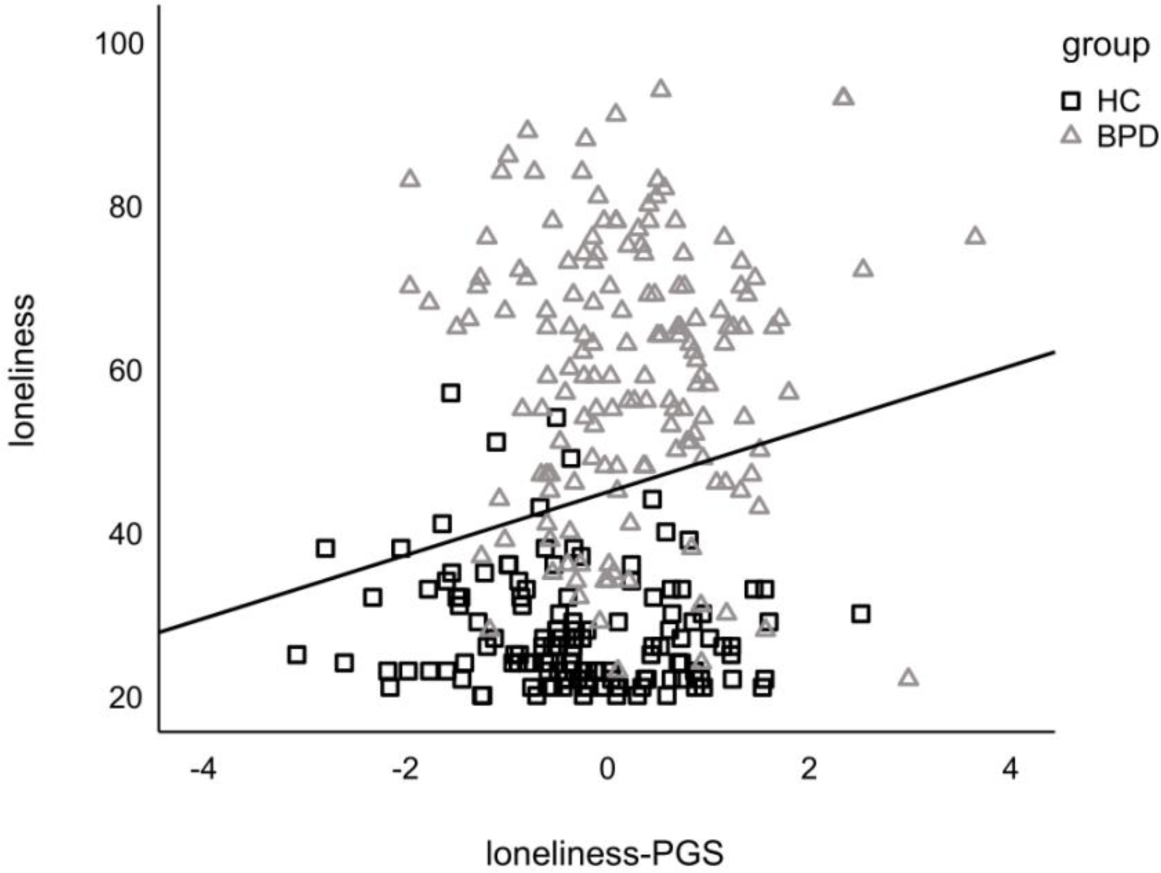
Association between the standardized loneliness-PGS and loneliness assessed with the ULS-R in HC and BPD.

### 3.4 Exploring loneliness-PGS as a potential modulating factor of the association between childhood traumatization and loneliness in BPD

To analyze the role of the genetic risk for loneliness as a vulnerability factor that might modulate the association of childhood traumatization and loneliness, moderation analysis was applied for the BPD group. The overall model was not significant, *F*(8, 135) = 1.712, *p* = .101, *R*² = .103. Results show that the loneliness-PGS did not moderate the effect between CTQ and loneliness, Δ*R*² = .004, *F*(1, 135) = 0.458, *p* = .500, [95% CI: –2.484; 5.067]. This did not change with the addition of the Neuroticism-PGS (overall *F*(9,134) = 1.506, *p* = .152, *R^2^* = .104; no interaction effect: *F*(1, 134) = 0.452, *p* = .503, Δ*R*² = .004, 95% CI: [-2.483; 5.038]; Table 1).

**Table 1.**
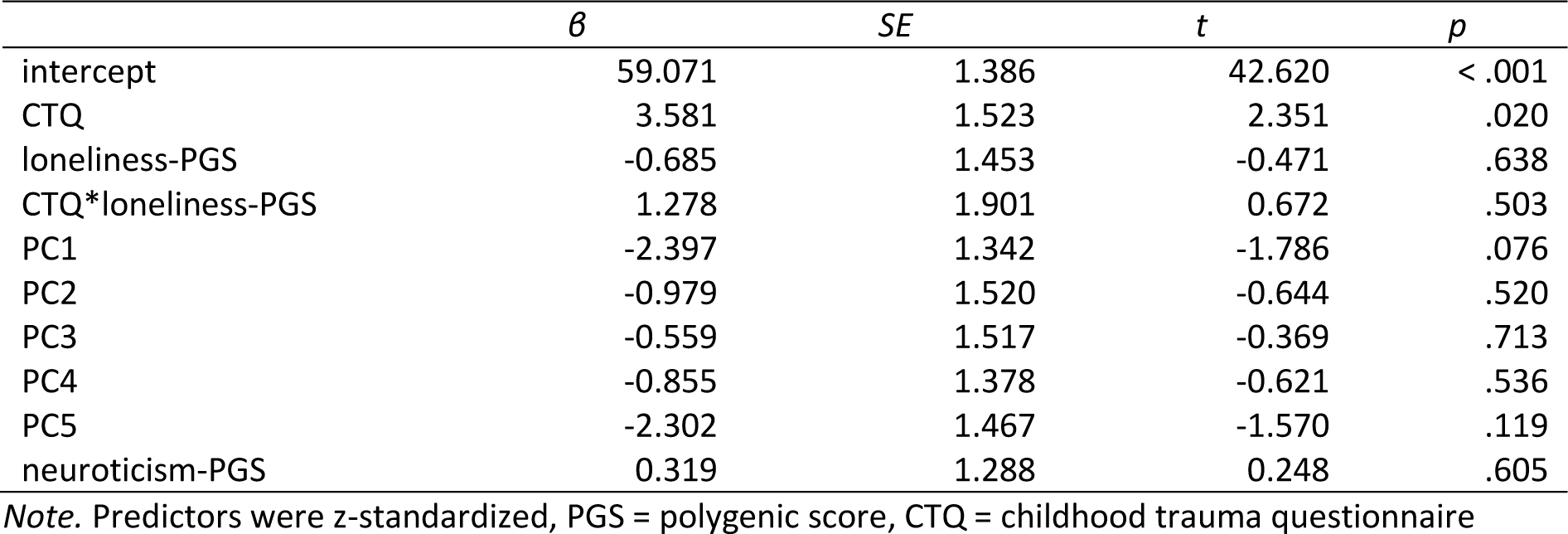
Prediction of loneliness by childhood maltreatment and Loneliness-PGS.

## 4 Discussion

The aims of the present study were to test a possible genetic overlap between loneliness and BPD, to examine whether a higher genetic risk for loneliness is associated with higher loneliness experienced by BPD patients, and to investigate whether the genetic risk for loneliness modulates the relationship of the severity of childhood maltreatment and experienced loneliness in BPD. Therefore, we examined genetic and self-report questionnaire data of patients with a clinical confirmed diagnosis of BPD and HC. We found evidence for a genetic overlap of BPD and loneliness, indicated by the genetic correlation of the two GWAS, and the higher loneliness-PGS in the BPD groups in both samples. In addition, a higher loneliness-PGS was associated with higher loneliness in the KFO-sample, but did not moderate the relationship between childhood maltreatment and loneliness. The associations remained even when controlling for the neuroticism-PGS, indicating that the genetic bridge between BPD and loneliness is partly but not only explained by a genetic propensity towards neuroticism as an anxious personality trait.

Our findings indicate that the genetic factors contributing to BPD risk and to variation in loneliness in the general population are partially shared via the observed genetic correlation of loneliness and BPD as well as the positive association of the loneliness-PGS with BPD case-control status. This is in line with former findings of a genetic association of borderline personality features and loneliness in a twin study [32]. Together with repeated findings on increased levels of loneliness and smaller social networks in BPD [19, 20], this finding underlines the relevance of loneliness in the context of BPD. The fact that a genetic correlation has already been shown for several other somatic and psychiatric diseases [38], supports prior research assuming loneliness as a transdiagnostically relevant risk factor [5–7].

In the combined sample of patients and controls from the KFO-sample, we found loneliness-PGS to be a positive predictor of self-reported loneliness, pointing towards the relevance of a genetic vulnerability for loneliness. The small effect size suggests that other components such as actual social isolation are important factors. That this association was not significant in the subgroups could be due to the fact that HCs and BPD represent extreme groups regarding experienced loneliness, showing reduced within-group but strong between-group variance. This suggests the need for further studies that enroll participants in both groups varying more broadly in the level of loneliness. While the current approach investigates the aggregate of the genetic association signal with loneliness and BPD, future studies in larger samples should examine which genes and pathways contribute to the genetic correlation. This might generate further insight in the underlying biological contribution to loneliness. In contrast to our hypothesis, the association of the severity of childhood maltreatment and reported loneliness was not moderated by the loneliness-PGS in the BPD group. While this suggests that that there are no interacting contributions of genetics and childhood maltreatment to subjective experienced loneliness, the lack of evidence may also have been caused by a lack of power due to the rather small sample. Although we have not found a significant interaction, future studies in larger samples should investigate whether subjects with an increased PGS for loneliness are especially vulnerable when additionally exposed to childhood maltreatment.

### 4.1 Limitations

The present study has some limitations. First, due to the overrepresentation of women with BPD in the health care system, our results are largely based on female subjects. In the Witt2017-sample, 92% of the BPD cases were female, and the KFO-sample consists of female participants only. Additionally, both samples were of central European ancestry. Therefore the generalisability is limited and replication in male or more balanced samples, and samples of other ancestries are needed.

Second, as already mentioned, our sample size was rather small for the investigation of the often small genetic effects. In this regard, we consider it a strength of the study, that the evidence for a shared genetic contribution to BPD and loneliness was replicated over different methods and two independent samples. However, especially for the more detailed analyses in the KFO-sample, there is need for studies replicating or extending those findings in larger samples. Larger samples would possibly allow to find effects that we could not confirm with our sample. Concurrently, larger GWAS samples, particularly for BPD, are warranted and would allow for more accurate estimations of genetic correlations, and more detailed biostatistical analyses of the shared genetics of loneliness and BPD, e.g. using methods taking both variants with equidirectional and opposing effects effect into account [61], or applying methods such as Mendelian randomisation to allow for inference of causality [62]. While prior studies on genes that are associated with loneliness reported enriched genetic signals for genes expressed in specific brain tissues in cortical and cerebellar regions [38], a conclusion on the genetic architecture is not possible based on our data, as we examined the aggregate of the genetic association signal. Larger GWAS samples for BPD would also allow methods such as local genetic correlations to be applied that are suitable to identify the genes and pathways shared between loneliness and BPD [63]. This could be helpful to further differentiate whether higher genetic risk directly affects feelings of loneliness or perceptions and behaviours that may lead to more loneliness. Thus, further research is needed to study the potentially complex interplay of genetic risk and e.g. personality dispositions such as rejection sensitivity and behaviors such as social withdrawal which might affect not only loneliness, but also the objective social isolation as indicated by smaller social networks in people with BPD [17–20].

Third, as a self-report questionnaire, the CTQ represents a retrospective assessment of childhood experiences rather than an objective description of the exposure and experiences of adverse childhood experiences [64]. That should be considered in the interpretation of effects of childhood maltreatment and emphasizes the need for studies with prospective designs. In addition, with the CTQ sum score we captured childhood maltreatment as a global measure, as a more detailed investigation of the interplay of different subtypes of maltreatment with the genetic propensity to loneliness was not possible due to the sample size. However, the type and timing of childhood maltreatment has been shown to influence its consequences [65] and should therefore be taken into account in future studies with larger samples, which would allow finer-grained analyses of different types of maltreatment such as emotional, physical and sexual abuse and neglect. Moreover, the measurement of the chronicity of loneliness might be the more appropriate tool to capture an association with genetic predispositions. Although the ULS-R is the most established instrument for measuring loneliness, originally conceptualized as a trait measure, it has been shown that most often it varies across time influenced by an individual’s current state instead of being exclusively a trait [66].

### 4.2 Conclusion

Despite the limitations mentioned above, our study is, as far as we know, the first study using genome-wide genetic data to link the polygenic propensity for loneliness to BPD, finding evidence for a higher genetic risk for loneliness in BPD compared to HC in two independent samples. Further studies and larger samples are needed to further dissect the genetic overlap, investigate possible effects of different types of childhood maltreatment interacting with the loneliness-PGS and address whether the association is specific for BPD or reflects a transdiagnostically relevant association. It is important to note that even though our findings have shown that genetic risk for loneliness explains some of the reported loneliness, this does not mean that it is not responsive to psychotherapeutic interventions. Therefore, our findings emphasize the importance of considering genetic risk when investigating the determinants of loneliness in BPD in order to tailor interventions like social skill training or the reappraisal of social interactions to the specific mechanism relevant to loneliness in people with BPD.

## Data Availability

The datasets presented in this article are not readily available because according to European law (GDPR), data containing potentially identifying or sensitive patient information are restricted; our data involving clinical participants are not freely available in the article, Supplementary Material, or in a public repository. Data are available upon reasonable request to the authors.

## 5 Acknowledgements

We thank all participants involved in the study. This project was funded the German Research Foundation (KFO-256 and GRK2350/3 – 324164820).

