## Supplementary Table for "Evidence for a shared genetic contribution to loneliness and Borderline Personality Disorder"

**Table S1**

*Number of SNPs included in PGS*

| | *loneliness* | | *neuroticism* | |
|---|---|---|---|---|
| **PT** | **sample 1** | **sample 2** | **sample 1** | **sample 2** |
| $5x10^{-8}$ | 11 | 12 | 128 | 118 |
| $1x10^{-6}$ | 36 | 37 | 275 | 248 |
| 0.0001 | 366 | 343 | 1183 | 1167 |
| 0.001 | 1363 | 1405 | 3056 | 3022 |
| 0.01 | 5730 | 6063 | 8696 | 8918 |
| 0.05 | 15,398 | 16,795 | 19,658 | 20,775 |
| 0.1 | 24,527 | 26,996 | 28,347 | 30,379 |
| 0.2 | 36,951 | 41,098 | 40,422 | 44,142 |
| 0.5 | 61,616 | 68,989 | 62,962 | 69,857 |
| 1 | 79,697 | 90,449 | 78,434 | 88,317 |

*Note*. Number of SNPs included for the calculation of polygenic scores (PGS) for loneliness and neuroticism at the different p-value thresholds, shown for both samples. SNP = single nucleotide polymorphism; PT = p-value threshold, PGS = polygenic score

**Table S2**

*Prediction of BPD case-control status by Loneliness-PGS in sample 1*

| ***Threshold*** | $R^2$ | ***P*** | ***Coefficient*** | ***Standard.Error*** | ***Num_SNP*** |
|---|---|---|---|---|---|
| *5,00E-08* | 0,02% | 0,485134 | 53,9157 | 77,2355 | 11 |
| *1,00E-06* | 0,24% | 0,020836 | 325,338 | 140,782 | 36 |
| *0,0001* | 0,43% | 0,002084 | 1892,59 | 614,891 | 366 |
| *0,001* | 0,43% | 0,0021 | 4194,05 | 1363,58 | 1363 |
| *0,01* | 1,19% | 3,83E-07 | 16524,5 | 3254,53 | 5730 |
| *0,05* | 1,72% | 1,13E-09 | 37523,7 | 6161,93 | 15398 |
| *0,1* | 2,02% | 4,43E-11 | 56111 | 8515,97 | 24527 |
| *0,2* | 2,26% | 3,21E-12 | 80217,4 | 11512,3 | 36951 |
| *0,5* | 2,27% | 2,73E-12 | 121418 | 17368,2 | 61616 |
| *1* | 2,14% | 1,11E-11 | 150398 | 22143,8 | 79697 |

*Note*. Threshold = P value threshold, for inclusion of SNPs, R2 = Variance explained by PGS, P = P value of association, Coefficient = Coefficient of PGS association, Standard.Error = Standard error of coefficient, Num_SNP = Number of included SNPs

**Table S3**

*Prediction of BPD case-control status by Loneliness-PGS in sample 2*

| *Threshold* | $R^2$ | *P* | *Coefficient* | *Standard.Error* | *Num_SNP* |
|---|---|---|---|---|---|
| *5,00E-08* | 0,52% | 0,184573 | 238,21 | 179,536 | 12 |
| *1,00E-06* | 0,02% | 0,780057 | 88,6995 | 317,641 | 37 |
| *0,0001* | 1,59% | 0,020926 | 3028,34 | 1311,36 | 343 |
| *0,001* | 1,06% | 0,058203 | 5771,3 | 3046,89 | 1405 |
| *0,01* | 4,26% | 0,000195 | 27890,4 | 7486,94 | 6063 |
| *0,05* | 5,39% | 2,99E-05 | 60046,1 | 14384,1 | 16795 |
| *0,1* | 6,60% | 4,41E-06 | 94635,6 | 20613,8 | 26996 |
| *0,2* | 6,46% | 5,26E-06 | 123476 | 27113,7 | 41098 |
| *0,5* | 5,32% | 3,34E-05 | 170799 | 41169,9 | 68989 |
| *1* | 5,27% | 3,57E-05 | 219538 | 53109 | 90449 |

*Note*. Threshold = P value threshold, for inclusion of SNPs, R2 = Variance explained by PGS, P = P value of association, Coefficient = Coefficient of PGS association, Standard.Error = Standard error of coefficient, Num_SNP = Number of included SNPs

**Table S4**

*Prediction of BPD case-control status by Neuroticism-PGS in sample 1*

| *Threshold* | $R^2$ | *P* | *Coefficient* | *Standard.Error* | *Num_SNP* |
|---|---|---|---|---|---|
| *5,00E-08* | 1,20% | 3,12E-07 | 0,976372 | 0,190851 | 128 |
| *1,00E-06* | 1,26% | 1,67E-07 | 1,70203 | 0,325235 | 275 |
| *0,0001* | 1,52% | 1,00E-08 | 5,06256 | 0,883501 | 1183 |
| *0,001* | 1,83% | 3,20E-10 | 10,8374 | 1,72329 | 3056 |
| *0,01* | 2,93% | 2,42E-15 | 28,3215 | 3,57706 | 8696 |
| *0,05* | 3,22% | 1,16E-16 | 52,9985 | 6,39506 | 19658 |
| *0,1* | 3,26% | 7,49E-17 | 69,6909 | 8,35724 | 28347 |
| *0,2* | 2,82% | 7,29E-15 | 85,4205 | 10,9805 | 40422 |
| *0,5* | 2,80% | 9,39E-15 | 125,074 | 16,1443 | 62962 |
| *1* | 2,68% | 3,32E-14 | 151,182 | 19,9308 | 78434 |

*Note*. Threshold = P value threshold, for inclusion of SNPs, R2 = Variance explained by PGS, P = P value of association, Coefficient = Coefficient of PGS association, Standard.Error = Standard error of coefficient, Num_SNP = Number of included SNPs

**Table S5**

*Prediction of BPD case-control status by Neuroticism-PGS in sample 2*

| *Threshold* | *$R^2$* | *P* | *Coefficient* | *Standard.Error* | *Num_SNP* |
|---|---|---|---|---|---|
| *5,00E-08* | 1,05% | 0,086132 | 0,858867 | 0,500458 | 118 |
| *1,00E-06* | 1,14% | 0,062194 | 1,42983 | 0,7667 | 248 |
| *0,0001* | 2,95% | 0,001887 | 6,24455 | 2,00948 | 1167 |
| *0,001* | 2,87% | 0,002042 | 11,6664 | 3,78271 | 3022 |
| *0,01* | 5,11% | 4,52E-05 | 31,9979 | 7,84412 | 8918 |
| *0,05* | 4,94% | 6,07E-05 | 59,0445 | 14,7243 | 20775 |
| *0,1* | 7,20% | 1,62E-06 | 97,3458 | 20,3 | 30379 |
| *0,2* | 5,93% | 1,21E-05 | 116,457 | 26,6188 | 44142 |
| *0,5* | 6,64% | 3,82E-06 | 185,185 | 40,0769 | 69857 |
| *1* | 6,68% | 3,58E-06 | 233,764 | 50,4433 | 88317 |

*Note*. Threshold = P value threshold, for inclusion of SNPs, R2 = Variance explained by PGS, P = P value of association, Coefficient = Coefficient of PGS association, Standard.Error = Standard error of coefficient, Num_SNP = Number of included SNPs

**Table S6**

*Prediction of BPD case-control status by Loneliness-PGS in sample 1, adjusted for Neuroticism-PGS*

| *Threshold* | *$R^2$* | *P* | *Coefficient* | *Standard.Error* | *Num_SNP* |
|---|---|---|---|---|---|
| *5,00E-08* | 0,00% | 0,928028 | 7,09112 | 78,5055 | 11 |
| *1,00E-06* | 0,05% | 0,285938 | 153,727 | 144,064 | 36 |
| *0,0001* | 0,07% | 0,214689 | 788,579 | 635,554 | 366 |
| *0,001* | 0,01% | 0,661796 | 633,814 | 1448,93 | 1363 |
| *0,01* | 0,19% | 0,036556 | 7364,58 | 3522,57 | 5730 |
| *0,05* | 0,34% | 0,005847 | 18823,3 | 6829,29 | 15398 |
| *0,1* | 0,47% | 0,001084 | 30925,7 | 9464,2 | 24527 |
| *0,2* | 0,62% | 0,000199 | 47475,4 | 12762,2 | 36951 |
| *0,5* | 0,62% | 0,000193 | 71895,5 | 19283,9 | 61616 |
| *1* | 0,55% | 0,000458 | 86204,7 | 24600,3 | 79697 |

*Note*. Threshold = P value threshold, for inclusion of SNPs, R2 = Variance explained by PGS, P = P value of association, Coefficient = Coefficient of PGS association, Standard.Error = Standard error of coefficient, Num_SNP = Number of included SNPs

**Table S7**

*Prediction of BPD case-control status by Loneliness-PGS in sample 2, adjusted for Neuroticism-PGS*

| ***Threshold*** | ***$R^2$*** | ***P*** | ***Coefficient*** | ***Standard.Error*** | ***Num_SNP*** |
|---|---|---|---|---|---|
| *5,00E-08* | 0,22% | 0,62118 | 160,864 | 325,517 | 12 |
| *1,00E-06* | 0,00% | 0,925435 | -42,5059 | 454,172 | 37 |
| *0,0001* | 0,67% | 0,157369 | 2102,7 | 1487,08 | 343 |
| *0,001* | 0,15% | 0,493603 | 2327,8 | 3400,28 | 1405 |
| *0,01* | 1,58% | 0,020136 | 18676,5 | 8037,06 | 6063 |
| *0,05* | 1,97% | 0,00903 | 40811,2 | 15631,1 | 16795 |
| *0,1* | 2,73% | 0,002126 | 68385,9 | 22260,9 | 26996 |
| *0,2* | 2,60% | 0,00264 | 88055,8 | 29285,1 | 41098 |
| *0,5* | 1,78% | 0,012332 | 111941 | 44731,7 | 68989 |
| *1* | 1,73% | 0,013757 | 142390 | 57799,1 | 90449 |

*Note*. Threshold = P value threshold, for inclusion of SNPs, R2 = Variance explained by PGS, P = P value of association, Coefficient = Coefficient of PGS association, Standard.Error = Standard error of coefficient, Num_SNP = Number of included SNPs
